# Antibody titers measured by commercial assays are correlated with neutralizing antibody titers calibrated by international standards

**DOI:** 10.1101/2021.07.16.21260618

**Authors:** Yu-An Kung, Chung-Guei Huang, Sheng-Yu Huang, Kuan-Ting Liu, Peng-Nien Huang, Kar-Yee Yu, Shu-Li Yang, Chia-Pei Chen, Ching-Yun Cheng, Yueh-Te Lin, Yen-Chin Liu, Guang-Wu Chen, Shin-Ru Shih

## Abstract

The World Health Organization (WHO) has highlighted the importance of an international standard (IS) for SARS-CoV-2 neutralizing antibody titer detection, with the aim of calibrating different diagnostic techniques. In this study, IS was applied to calibrate neutralizing antibody titers (IU/mL) and binding antibody titers (BAU/mL) in response to SARS-CoV-2 vaccines. Serum samples were collected from participants receiving the Moderna (n = 20) and Pfizer (n = 20) vaccines at three time points: pre-vaccination, after one dose, and after two doses. We obtained geometric mean titers of 1404.16 and 928.75 IU/mL for neutralizing antibodies after two doses of the Moderna and Pfizer vaccines, respectively. These values provide an important baseline for vaccine development and the implementation of non-inferiority trials. We also compared three commercially available kits from Roche, Abbott, and MeDiPro for the detection of COVID-19 antibodies based on binding affinity to S1 and/or RBD. Our results demonstrated that antibody titers measured by commercial assays are highly correlated with neutralizing antibody titers calibrated by IS.

## Introduction

Identifying immune correlates of protection against SARS-CoV-2 infection is challenging. Neutralizing antibody titer is not the only determinant of vaccine efficacy; however, the neutralization level is highly predictive of immune protection (1-3). Moreover, the detection of neutralizing antibody titers is feasible in many laboratories. Several methods have been developed to measure neutralizing antibody titers in convalescent serum or vaccinated serum, such as plaque reduction assays, focus reduction assays, microneutralization assays using real viruses, and pseudovirus assays (4, 5). These assays differ substantially, including variation in protocols for similar assay types and variation among laboratories (6, 7).

For calibration, the establishment of international standards (IS) for SARS-CoV-2 neutralizing antibody titer detection is a key goal of the World Health Organization (WHO). Various standards, such as 20/130 and 20/136 (provided by National Institute for Biological Standards and Control [NIBSC]), are widely used to establish a baseline for comparing neutralizing antibody titers from different datasets (different laboratories, protocols, assays, etc.) (6). An IS is based on pooled human plasma from convalescent patients (8). For different pooled cohorts, different standard sera are obtained, each with predetermined international units (IU) for the conversion of neutralizing antibody titers, enabling immunogenicity characteristics to be correctly compared across laboratories and vaccine developers. Although many laboratories have applied IU to present neutralizing antibody titers in the serum of patients with COVID-19 or vaccinated individuals, reference data are insufficient.

Furthermore, neutralization tests using real viruses at biosafety level 3 (BSL-3) laboratories are laborious and time-consuming. Binding assays based on the anti-spike protein or anti-receptor binding domain (RBD) are widely used to measure neutralizing antibodies (9-11). However, the correlation between antibody titers from binding assays (binding antibody unit, BAU) and neutralizing titers (IU) has not been clearly established.

In this study, 120 serum samples from 40 total subjects were analyzed (20 subjects receiving Moderna vaccines and 20 subjects receiving the Pfizer vaccines) before vaccination, after the first dose, and after the second dose to estimate the correlation between titers estimated by a microneutralization assay using real viruses in a BSL-3 laboratory (IU/mL) and antibody titers measured by anti-S1 and anti-RBD enzyme-linked immunosorbent assay (ELISA) (BAU/mL). Several promising vaccine candidates have undergone different phases of clinical trials (12). Our results provide useful information for comparisons of neutralizing antibody titers with those of other widely used vaccines and for non-inferiority trials.

## Materials and Methods

### Serum samples collection

A total of 120 COVID-19 vaccinated sera were purchased from Access Biologicals (Vista, CA, USA). The serum samples were collected and followed by protocol SDP-003, *Human Biological Specimens Collection*, data September 22, 2017 and the qualifications of Principle Investigator (Robert Pyrtle, M.D.) were reviewed and approved by Diagnostics Investigational Review Board (Cummaquid, Massachusetts, USA). The protocol SDP-003 will expire on May 3, 2022. The collection dates of the sera were between February 25, 2021 to April 29, 2021. The serum from 20 individuals vaccinated with Moderna mRNA-1273 and 20 individuals vaccinated with Pfizer BNT162b2 were collected before vaccination, after the first dose, and after the second dose. The ages of the vaccinated participants are between 22 to 69 years old.

### Cell culture and virus

African green monkey kidney (Vero E6) cells (CRL-1586) were purchased from the American Type Culture Collection (ATCC, Bethesda, MD, USA) and maintained in Dulbecco’s modified Eagle’s medium (DMEM; Gibco, Waltham, MA, USA) containing 10% fetal bovine serum (FBS; Gibco) at 37°C. Severe acute respiratory syndrome coronavirus 2 isolate SARS-CoV-2/human/TWN/CGMH-CGU-01/2020 was used in the live virus microneutralization assay.

### Live virus microneutralization assay

Vero E6 cells (2 × 10^4^ cells per well) were seeded in a 96-well plate and incubated at 37°C for 24 h. The medium was replaced with 100 μL of fresh DMEM containing 2% FBS. The live virus microneutralization assay was performed in a BSL-3 laboratory using the SARS-CoV-2/human/TWN/CGMH-CGU-01/2020 strain. All serum samples were heat-inactivated at 56°C for 30 min and then 2-fold serially diluted in DMEM (Gibco) without FBS. From a starting dilution of 1:8 for each sample, ten 2-fold dilutions were performed for a final dilution of 1:8192. Each serum sample was incubated with 100 50% tissue culture infectious doses (100 TCID50) of SARS-CoV-2 at 37°C for 1 h prior to infection with Vero E6 cells. Add 100 μL of the virus-serum mixtures at each dilution to a 96-well plate containing the confluent Vero E6 monolayer. After infected cells were incubated at 37°C for 5 days, they were fixed with 10% formaldehyde and stained with crystal violet. The neutralization titer was calculated as the logarithm of the 50% end point using the Reed–Muench method based on the presence or absence of cytopathic effects. Each serum sample was tested in four replicates. Geometric mean titers (GMTs) were calculated with 95% confidence intervals (CIs) using GraphPad Prism version 8 (GraphPad Software, Inc., CA, USA).

### ELISA

For indirect ELISA, a 96-well plate was coated with 2 μg/mL S1, RBD, and N protein (Sino Biological, Beijing, China) diluted in phosphate-buffered saline and incubated overnight at 4°C. Each well was blocked with 300 μL of StartingBlock™ T20 blocking buffer (Thermo Fisher Scientific, Waltham, MA, USA) for 1 h at 37°C. Serum from vaccinated donors or NIBSC 20/136 standard was diluted in blocking buffer, and 100 μL of the sample was added to a 96-well plate, followed by incubation for 1 h at 37°C. After washing, horseradish peroxidase (HRP)-tagged anti-human antibodies (Abcam, Cambridge, UK), diluted 1:10,000 in blocking buffer, were added to the wells (100 μL/well), and the plate was incubated for 1 h at 37°C. Samples with N antibodies were incubated for 30 min at 37°C. The chromogenic reagent 3,3,5,5-tetramethylbenzidine (TMB) was mixed with an equal volume of Color A and B (R&D Systems, Minneapolis, MN, USA). The TMB reaction time for S1 and RBD ELISA was 5 min and for N protein ELISA was 10 min. After the reaction, stop solution (R&D Systems) was added to the wells, and the absorbance was measured immediately at 450 nm using a Synergy 2 Microplate Reader (Bio-Tek, Winooski, VT, USA).

### Serologic assay

Each serum sample was analyzed by the MeDiPro SARS-CoV-2 antibody ELISA, Roche Elecsys^®^ Anti-SARS-CoV-2 S assay, and Abbott AdviseDx SARS-CoV-2 IgG II assay, according to the manufacturers’ instructions. The MeDiPro SARS-CoV-2 antibody ELISA detected antibodies against S1 and RBD, and values <34.47 IU/mL were considered negative. The electrochemiluminescence immunoassay (ECLIA) (i.e., Roche Elecsys Anti-SARS-CoV-2 S assay) was used for the detection of antibodies against the RBD of S protein; <0.80 U/mL was considered negative and ≥0.80 U/mL was considered positive for anti-SARS-CoV-2 S protein. The Abbott AdviseDx SARS-CoV-2 IgG II assay is a chemiluminescent microparticle immunoassay (CMIA) for the detection of IgG antibodies to the RBD of S protein; the cut-off value was 50.0 AU/mL.

### WHO international standard unit (IU) conversion

WHO IS sera (20/130, 20/136, and 20/268) were obtained from NIBSC. The 50% neutralization titer (NT_50_) values for WHO IS sera were determined by a live virus microneutralization assay (Supplementary Table 1). Each standard serum sample was tested in duplicate, except 20/130.

### Statistical analysis

Statistical analyses were performed using GraphPad Prism version 8 (GraphPad Software, Inc., CA, USA). Pearson’s correlation coefficients (*r*) were used to determine the correlation between the titers obtained by the different serological assays and the live SARS-CoV-2 NT assay. Statistical significance was set at *P* < 0.05.

## Results

The neutralizing antibody titer is important for evaluating protection against viral infection after vaccination. Dynamic neutralizing antibody titers were observed in individuals who had been fully vaccinated. In detail, we obtained serum samples from 20 individuals vaccinated with Moderna mRNA-1273 and 20 individuals vaccinated with Pfizer BNT162b2. For each of these 40 individuals, serum samples were collected before the first dose, 24 days after the first dose from Moderna or 14 days after the first dose from Pfizer, and 14 days after the second dose. A total of 120 serum samples were tested to determine neutralizing antibody titers by a live SARS-CoV-2 virus microneutralization assay. We obtained the NT_50_ values that represent 50% protection against SARS-CoV-2-induced cell death. As expected, the neutralizing antibody titers increased after the first and second doses of the Moderna and Pfizer vaccines (Figure 1A). The NT_50_ values for the WHO IS for anti-SARS-CoV-2 immunoglobulin (obtained from NIBSC) were also determined by a live virus neutralization assay (Supplementary Table 1), with linear regression defining the conversion of NT_50_ values to IU/mL, as shown in Figure 1B. The GMTs are also shown in Figure 1, illustrating the observed increases. The GMT increased from 157.6 to 1404.16 IU/mL and from 348.83 to 928.75 IU/mL for Moderna and Pfizer vaccines, respectively. These GMT values provide a reference for non-inferiority tests of candidate vaccines.

**Figure 1.**
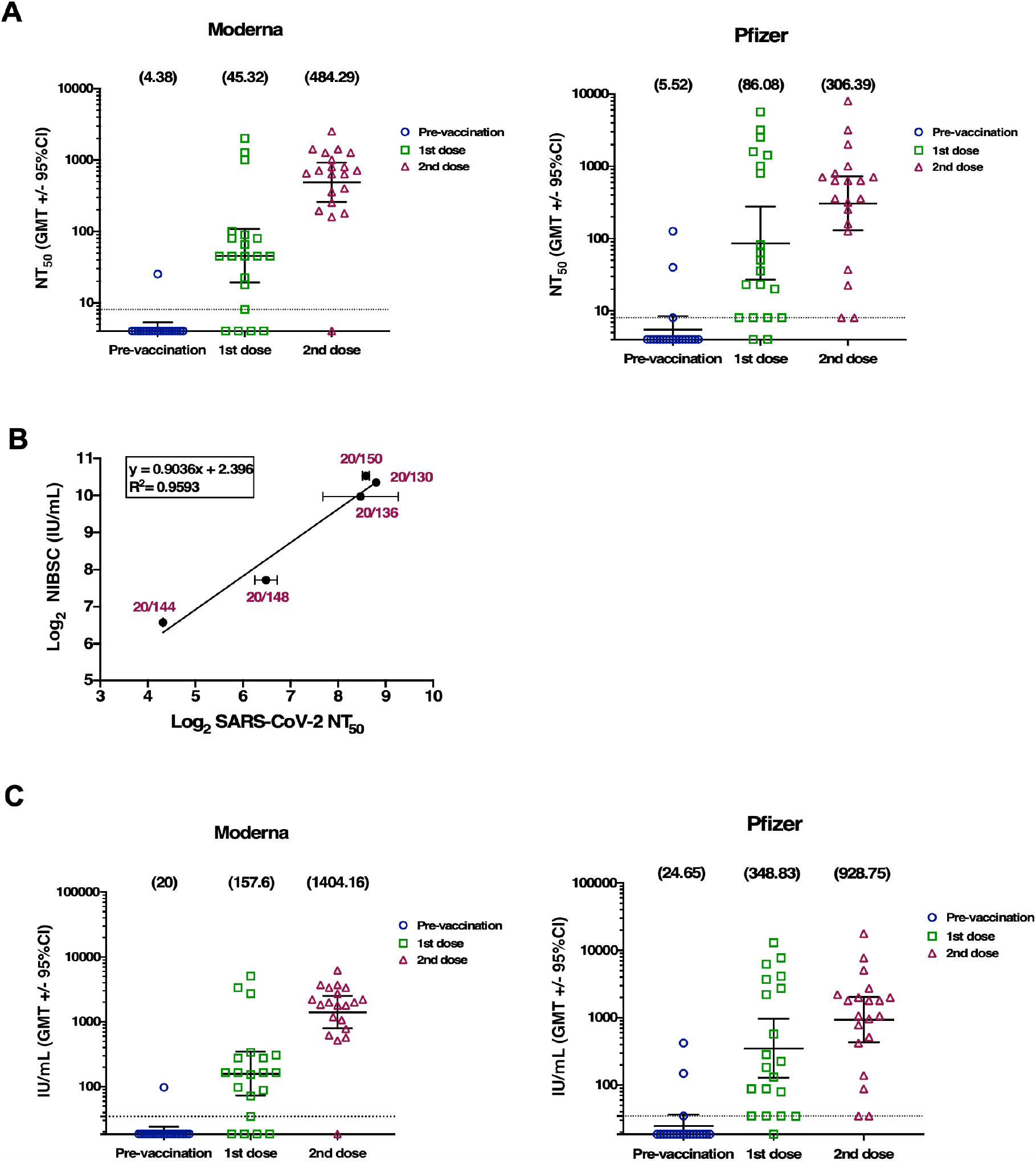
Neutralization serum titers in vaccinated individuals. **(A)** NT_50_ values for 120 serum samples from 20 recipients of Moderna mRNA-1273 and 20 recipients of Pfizer BNT162b2. **(B)** The calibration curve (standard curve) used for the conversion of NT_50_ values to the international standard units (IU/mL). Presented are the results from an experiment in technical duplicate and error bars show the SD. **(C)** The IU value for 120 serum samples (20 for the Moderna mRNA-1273 group and 20 for the Pfizer BNT162b2 group). The geometric mean titers (GMTs) with 95% CI are shown pre-vaccination, after the first dose, and after the second dose.

Although the live virus neutralization assay is the gold standard for determining neutralizing antibody titers, the operation is time-consuming and requires a BSL-3 laboratory. We developed binding assays to detect anti-S1, anti-RBD, and anti-N antibodies, followed by NIBSC 20/136 (WHO IS) for conversion to BAU/mL (Figure 2). Increases in anti-S1 and anti-RBD antibodies were observed after the first and second doses (Figure 2A and B). Interestingly, anti-N antibodies were detected in a few individuals pre- and post-vaccination, suggesting that the individuals had COVID-19 before or after vaccination.

**Figure 2.**
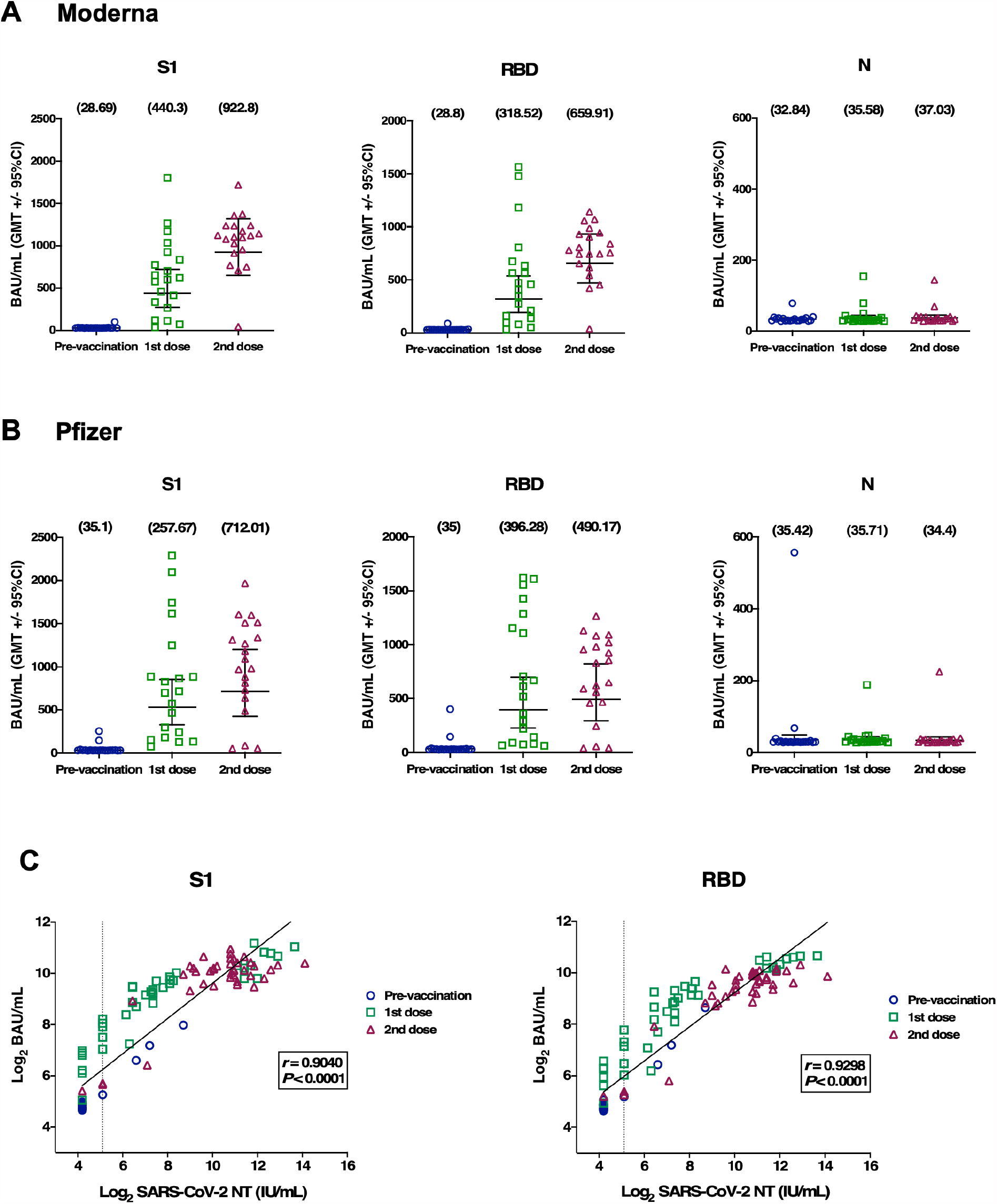
Antibody response in 120 serum samples from vaccinated individuals. The responses of antibodies against S1, RBD, and N protein were detected in 120 serum samples from 20 individuals receiving Moderna mRNA-1273 (**A**) and 20 individuals receiving Pfizer BNT162b2 vaccines (**B**). The geometric mean titers (GMTs) with 95% CI are shown pre-vaccination, after the first dose, and after the second dose. (**C**) Correlation between the live virus neutralization titer (IU/mL) and antibody binding unit (BAU/mL) in 120 serum samples. Vertical dashed lines indicate the limit of detection (NT = 34.47). The Pearson’s correlation coefficients (*r*) are provided for S1 or RBD antibody responses to live virus neutralization titer (IU/mL).

To evaluate whether our binding assay based on anti-S1 or anti-RBD in BAU/mL reflects neutralizing antibody titers in IU/mL, the correlation between the two values was determined, as shown in Figure 2C. Both the S1 and RBD antibody titers were highly correlated with the NT titers (*r* = 0.9040 and 0.9298, respectively), suggesting that the binding assays based on anti-S1 or RBD can be used as surrogates to measure neutralizing antibodies.

All 120 serum samples were tested by commercial serological assays, including the MeDiPro SARS-CoV-2 antibody ELISA, Roche Elecsys^®^ Anti-SARS-CoV-2 S, and Abbott AdviseDx SARS-CoV-2 IgG II assay. Roche and Abbott serological assays are widely used in clinical laboratories to detect SARS-CoV-2 antibodies worldwide. They detect the antibody against the RBD of the S antigen, yielding qualitative and semi-quantitative results. MeDiPro is a Taiwan FDA-approved kit for quantifying S1- and RBD-binding antibodies; it assumes that data for S1 and RBD fusion proteins can accurately predict the NT titer. We used real NT titers (IU/mL) from BSL-3 as a standard to assess whether these serological assays reflect the neutralization titers based on the detection of antibodies against S1 and/or RBD. The highest correlation was observed between the titers obtained by the MeDiPro and the live SARS-CoV-2 NT assays (*r* = 0.9111) (Figure 3A). Roche and Abbott RBD antibody titers also had good correlation coefficients of 0.7294 and 0.8466, respectively (Figure 3B and C). With respect to the live virus microneutralization assays, the Roche and Abbott serological assays can correctly detect RBD antibodies in all 77 NT-positive samples (Table 1), whereas MeDiPro produced six negative results. Among these six negative results, the NT titer of four samples is equal to 34.47 IU/mL, which is the limit of the detection in MeDiPro assay. Therefore, both the Roche and Abbot serological assays have 100% (95% Cl, 95.3%-100%) sensitivity, followed by 92.2% (95% Cl, 84.0%-96.4%) sensitivity for MeDiPro. However, MeDiPro can distinguish a greater proportion of NT-negative samples (40/43) than those of Roche (24/43) and Abbott (25/43). In terms of specificity, MeDiPro achieved a high specificity of 93% (95% Cl, 81.4%-97.6%), which was much higher than of the Roche (55.8% [95% Cl, 41.1%-69.6%]) and Abbott assays (58.1% [95% Cl, 43.3%-71.6%]). Our results suggest that the MeDiPro SARS-CoV-2 neutralization antibody assay is an effective option for detecting SARS-Cov-2 neutralizing antibodies without requiring a live virus neutralization assay at a BSL-3 laboratory.

**Table 1.** Comparison of commercial serological assays with SARS-CoV-2 neutralizing antibody titer.

| Total samples (120) | MeDiPro |  | Roche |  | Abbott |  |
| --- | --- | --- | --- | --- | --- | --- |
| Real NT | Positive | <sup>b</sup>Negative | Positive | <sup>c</sup>Negative | Positive | <sup>d</sup>Negative |
| Positive 77 | 71 | <sup>ψ</sup>6 | 77 | 0 | 77 | 0 |
| <sup>a</sup>Negative 43 | 3 | 40 | 19 | 24 | 18 | 25 |
| Sensitivity=TP/(TP+FN) | 92.2% (84.0%-96.4%) |  | 100% (95.3%-100%) |  | 100% (95.3%-100%) |  |
| Specificity=TN/(TN+FP) | 93.0% (81.4%-97.6%) |  | 55.8% (41.1%-69.6%) |  | 58.1% (43.3%-71.6%) |  |
| PPV=TP/(TP+FP) | 95.9% (85.8%-98.9%) |  | 80.2% (71.1%-87.0%) |  | 81.1% (72.0%-87.7%) |  |
| NPV=TN/(TN+FN) | 87.0% (74.3%-93.9%) |  | 100% (86.2%-100%) |  | 100.0% (86.7%-100%) |  |

TP, true positive; FP, false positive; TN, true negative; FN, false negative; PPV,
positive predictive value; NPV, negative predictive value
<sup>a, b</sup> Negative < 34.37 IU/mL
<sup>c</sup> Negative < 0.80 U/mL
<sup>d</sup> Negative < 50.0 AU/mL
<sup>ψ</sup> Among the six negative samples, four samples have a neutralizing antibody titer of
34.37 IU/mL.

**Figure 3.**
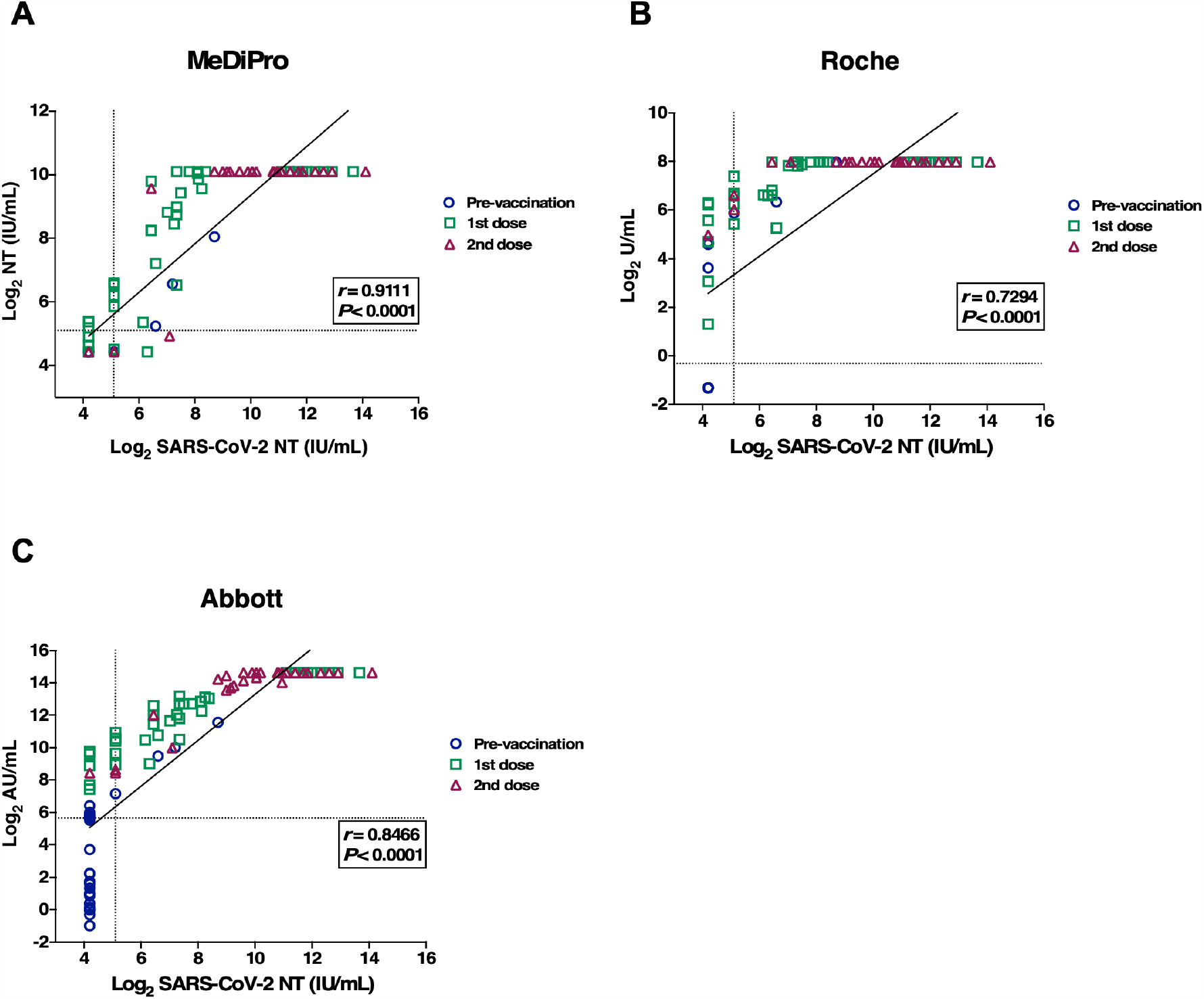
Correlation analysis of commercial serological assays against SARS-CoV-2 neutralizing antibody titers. Correlations between titers obtained by MeDiPro SARS-CoV-2 antibody ELISA (**A**), Roche Elecsys^®^ Anti-SARS-CoV-2 S assay (**B**), and Abbott AdviseDx SARS-CoV-2 IgG II assay (**C**) and live SARS-CoV-2 neutralization titers (IU/mL). The vertical dashed line indicates the limit of detection (NT = 34.47 IU/mL). The horizontal dashed lines indicate the cut-off values for MeDiPro (34.47 IU/mL), Roche (0.80 U/mL), and Abbott (50.0 AU/mL). Correlations were performed in the Pearson’s correlation coefficients (*r*).

## Discussion

Antibodies increase gradually within a few weeks after vaccination, and the timespan may vary among individuals (13, 14). Therefore, it is necessary to test neutralizing antibodies to determine whether protective antibodies will be elevated after vaccination. Vaccinated individuals may still have to take measures to avoid infection. Accordingly, such assays are very important for protecting vaccinated individuals as well as for the control and prevention of epidemics (15).

Anti-N antibodies may reveal whether vaccinated individuals were infected by the virus before or after the vaccination dose (16). For example, as shown in Figure 2B, one individual who received the Pfizer vaccine showed a dramatic increase in N to 556 BAU/mL. S1 for the same individual, after the first and second doses, were 2,094 and 1,240 BAU/mL, respectively. In the same individual, after the first and second doses, the RBD levels were 1,618 and 927 BAU/mL, respectively. It has been reported that prior infection with SARS-CoV-2 may boost B cells and significantly elevate antibody production (17, 18).

Roche and Abbott Covid-19 diagnostic kits are being used extensively in clinical laboratories (19-21). They are semi-quantitative and were originally designed to confirm an infected case. In this study, we utilized standard sera to develop an approach that utilizes these two kits to quantitate antibody titers after vaccination. The Pearson’s correlation coefficients (*r*) between these antibody titers and neutralization antibody titers were 0.7294 and 0.8466, respectively. Titers obtained by MeDiPro, designed to detect neutralizing antibody titers, were highly correlated with titers obtained by live SARS-CoV-2 NT assays (*r* = 0.9111).

IS was used to obtain neutralizing antibody titers in IU/mL and binding antibody titers in BAU/mL. We obtained GMTs of 1404.16 and 928.75 for neutralizing antibodies in serum samples from recipients of Moderna and Pfizer vaccines (n = 20 each) after two doses, respectively. These estimates may provide important information for vaccine developers implementing non-inferiority tests. Moreover, we compared three commercially available kits used for the detection of COVID-19 antibodies based on binding affinities to S1 and/or RBD. Our results demonstrated that antibody titers measured by commercial assays are strongly correlated with neutralizing antibody titers via IS calibration.

## Data Availability

The data that support the findings of this study are available from the corresponding author, upon reasonable request.

## Acknowledgements

This work was financially supported by the Research Center for Emerging Viral Infections from The Featured Areas Research Center Program within the framework of the Higher Education Sprout Project by the Ministry of Education (MOE), Taiwan, the Ministry of Science and Technology (MOST), Taiwan (MOST 109-2634-F-182-001 and 109-2221-E-182-043-MY2), the Research Center for Epidemic Prevention Science by the MOST, Taiwan (MOST 109-2327-B-182-002), the Chang Gung Memorial Hospital (grant number BMRP367), and the National Institutes of Health USA grant U01 AI151698 for the United World Antiviral Research Network (UWARN).

## Author contributions

C.G.H, G.W.C. and S.R.S. designed the experiments. Y.A.K., S.Y.H., P.N.H., Y.T.L. and Y.C.L. conducted the live virus neutralization assay at BSL3 facility. K.T.L., K.Y.Y., S.L.Y., C.P.C. and C.Y.C conducted the serological assay. Y.A.K., S.Y.H.,K.T.L. and C.G.H. analyzed the data. Y.A.K., K.T.L., G.W.C. and S.R.S. wrote the manuscript.

## Declaration

MeDiPro SARS-CoV-2 antibody ELISA was technology transferred from Research Center for Emerging Viral Infections, Chang Gung University, Taiwan. We herewith declare that MeDiPro, Roche, and Abbott did not financially support any research in Research Center for Emerging Viral Infections, Chang Gung University and Chang Gung Memorial Hospital, Taiwan.

## Supplementary information

**Supplementary Table 1.** The NT_50_ values of WHO international standard sera.

|  | Research<br>Reagent | WHO<br>International<br>Standard | WHO Reference Panel |  |  |  |  |
| --- | --- | --- | --- | --- | --- | --- | --- |
|  |  |  | NIBSC 20/268 |  |  |  |  |
| NIBSC Code | NIBSC<br>20/130 | NIBSC<br>20/136 | High<br>20/150 | Mid<br>20/148 | low S,<br>high N<br>20/144 | Low<br>20/140 | NC<br>20/142 |
| Neutralizing Ab<br>(IU/mL) | 1300 | 1000 | 1473 | 210 | 95 | 44 | — |
| Neutralizing Ab<br>(NT<sub>50</sub>) Test-1 | 446.8 | 223.89 | 398.11 | 79.43 | 20 | <20 | <20 |
| Neutralizing Ab<br>(NT<sub>50</sub>) Test-2 |  | 486.97 | 370.88 | 100 | 19.95 | <20 | <20 |

NIBSC, National Institute for Biological Standards and Control; NT<sub>50</sub>, 50%
neutralization titer

## References

1. Khoury DS, Cromer D, Reynaldi A, Schlub TE, Wheatley AK, Juno JA, Subbarao K, Kent SJ, Triccas JA, Davenport MP. 2021. Neutralizing antibody levels are highly predictive of immune protection from symptomatic SARS-CoV-2 infection. Nat Med doi:10.1038/s41591-021-01377-8.

2. Chia WN, Zhu F, Ong SWX, Young BE, Fong SW, Le Bert N, Tan CW, Tiu C, Zhang J, Tan SY, Pada S, Chan YH, Tham CYL, Kunasegaran K, Chen MI, Low JGH, Leo YS, Renia L, Bertoletti A, Ng LFP, Lye DC, Wang LF. 2021. Dynamics of SARS-CoV-2 neutralising antibody responses and duration of immunity: a longitudinal study. Lancet Microbe 2:e240–e249.

3. Earle KA, Ambrosino DM, Fiore-Gartland A, Goldblatt D, Gilbert PB, Siber GR, Dull P, Plotkin SA. 2021. Evidence for antibody as a protective correlate for COVID-19 vaccines. Vaccine 39:4423–4428.

4. Bewley KR, Coombes NS, Gagnon L, McInroy L, Baker N, Shaik I, St-Jean JR, St-Amant N, Buttigieg KR, Humphries HE, Godwin KJ, Brunt E, Allen L, Leung S, Brown PJ, Penn EJ, Thomas K, Kulnis G, Hallis B, Carroll M, Funnell S, Charlton S. 2021. Quantification of SARS-CoV-2 neutralizing antibody by wild-type plaque reduction neutralization, microneutralization and pseudotyped virus neutralization assays. Nat Protoc 16:3114–3140.

5. Supasa P, Zhou D, Dejnirattisai W, Liu C, Mentzer AJ, Ginn HM, Zhao Y, Duyvesteyn HME, Nutalai R, Tuekprakhon A, Wang B, Paesen GC, Slon-Campos J, Lopez-Camacho C, Hallis B, Coombes N, Bewley KR, Charlton S, Walter TS, Barnes E, Dunachie SJ, Skelly D, Lumley SF, Baker N, Shaik I, Humphries HE, Godwin K, Gent N, Sienkiewicz A, Dold C, Levin R, Dong T, Pollard AJ, Knight JC, Klenerman P, Crook D, Lambe T, Clutterbuck E, Bibi S, Flaxman A, Bittaye M, Belij-Rammerstorfer S, Gilbert S, Hall DR, Williams MA, Paterson NG, James W, Carroll MW, Fry EE, Mongkolsapaya J, et al. 2021. Reduced neutralization of SARS-CoV-2 B.1.1.7 variant by convalescent and vaccine sera. Cell 184:2201–2211 e7.

6. WHO/BS.2020.2403. 2020. Establishment of the WHO International Standard and Reference Panel for anti-SARS-CoV-2 antibody.

7. WHO/BS.2020.2402. 2020. Collaborative Study for the Establishment of a WHO International Standard for SARS-CoV-2 RNA.

8. Kristiansen PA, Page M, Bernasconi V, Mattiuzzo G, Dull P, Makar K, Plotkin S, Knezevic I. 2021. WHO International Standard for anti-SARS-CoV-2 immunoglobulin. Lancet 397:1347–1348.

9. Tan CW, Chia WN, Qin X, Liu P, Chen MI, Tiu C, Hu Z, Chen VC, Young BE, Sia WR, Tan YJ, Foo R, Yi Y, Lye DC, Anderson DE, Wang LF. 2020. A SARS-CoV-2 surrogate virus neutralization test based on antibody-mediated blockage of ACE2-spike protein-protein interaction. Nat Biotechnol 38:1073–1078.

10. Patel EU, Bloch EM, Clarke W, Hsieh YH, Boon D, Eby Y, Fernandez RE, Baker OR, Keruly M, Kirby CS, Klock E, Littlefield K, Miller J, Schmidt HA, Sullivan P, Piwowar-Manning E, Shrestha R, Redd AD, Rothman RE, Sullivan D, Shoham S, Casadevall A, Quinn TC, Pekosz A, Tobian AAR, Laeyendecker O. 2021. Comparative Performance of Five Commercially Available Serologic Assays To Detect Antibodies to SARS-CoV-2 and Identify Individuals with High Neutralizing Titers. J Clin Microbiol 59.

11. Huynh A, Arnold DM, Smith JW, Moore JC, Zhang A, Chagla Z, Harvey BJ, Stacey HD, Ang JC, Clare R, Ivetic N, Chetty VT, Bowdish DME, Miller MS, Kelton JG, Nazy I. 2021. Characteristics of Anti-SARS-CoV-2 Antibodies in Recovered COVID-19 Subjects. Viruses 13.

12. Kyriakidis NC, Lopez-Cortes A, Gonzalez EV, Grimaldos AB, Prado EO. 2021. SARS-CoV-2 vaccines strategies: a comprehensive review of phase 3 candidates. NPJ Vaccines 6:28.

13. Goel RR, Apostolidis SA, Painter MM, Mathew D, Pattekar A, Kuthuru O, Gouma S, Hicks P, Meng W, Rosenfeld AM, Dysinger S, Lundgreen KA, Kuri-Cervantes L, Adamski S, Hicks A, Korte S, Oldridge DA, Baxter AE, Giles JR, Weirick M E, McAllister CM, Dougherty J, Long S, D’Andrea K, Hamilton JT, Betts MR, Luning Prak ET, Bates P, Hensley SE, Greenplate AR, Wherry EJ. 2021. Distinct antibody and memory B cell responses in SARS-CoV-2 naive and recovered individuals following mRNA vaccination. Sci Immunol 6.

14. Pollard AJ, Bijker EM. 2021. A guide to vaccinology: from basic principles to new developments. Nat Rev Immunol 21:83–100.

15. Bartsch SM, O’Shea KJ, Ferguson MC, Bottazzi ME, Wedlock PT, Strych U, McKinnell JA, Siegmund SS, Cox SN, Hotez PJ, Lee BY. 2020. Vaccine Efficacy Needed for a COVID-19 Coronavirus Vaccine to Prevent or Stop an Epidemic as the Sole Intervention. Am J Prev Med 59:493–503.

16. Diao B, Wen K, Zhang J, Chen J, Han C, Chen Y, Wang S, Deng G, Zhou H, Wu Y. 2021. Accuracy of a nucleocapsid protein antigen rapid test in the diagnosis of SARS-CoV-2 infection. Clin Microbiol Infect 27:289 e1–289 e4.

17. Reynolds CJ, Pade C, Gibbons JM, Butler DK, Otter AD, Menacho K, Fontana M, Smit A, Sackville-West JE, Cutino-Moguel T, Maini MK, Chain B, Noursadeghi M, Network UKCIC, Brooks T, Semper A, Manisty C, Treibel TA, Moon JC, Investigators UKC, Valdes AM, McKnight A, Altmann DM, Boyton R. 2021. Prior SARS-CoV-2 infection rescues B and T cell responses to variants after first vaccine dose. Science doi:10.1126/science.abh1282.

18. Anichini G, Terrosi C, Gandolfo C, Gori Savellini G, Fabrizi S, Miceli GB, Cusi MG. 2021. SARS-CoV-2 Antibody Response in Persons with Past Natural Infection. N Engl J Med 385:90–92.

19. Tan SS, Saw S, Chew KL, Wang C, Pajarillaga A, Khoo C, Wang W, Ali ZM, Yang Z, Chan YH, Tambyah P, Jureen R, Sethi SK. 2021. Comparative Clinical Evaluation of the Roche Elecsys and Abbott Severe Acute Respiratory Syndrome Coronavirus 2 (SARS-CoV-2) Serology Assays for Coronavirus Disease 2019 (COVID-19). Arch Pathol Lab Med 145:32–38.

20. Harley K, Gunsolus IL. 2020. Comparison of the Clinical Performances of the Abbott Alinity IgG, Abbott Architect IgM, and Roche Elecsys Total SARS-CoV-2 Antibody Assays. J Clin Microbiol 59.

21. Speletas M, Kyritsi MA, Vontas A, Theodoridou A, Chrysanthidis T, Hatzianastasiou S, Petinaki E, Hadjichristodoulou C. 2020. Evaluation of Two Chemiluminescent and Three ELISA Immunoassays for the Detection of SARS-CoV-2 IgG Antibodies: Implications for Disease Diagnosis and Patients’ Management. Front Immunol 11:609242.

